# Efficacy and safety of masitinib in amyotrophic lateral sclerosis patients prior to loss of functionality: A subgroup analysis optimizing the benefit-risk profile of masitinib

**DOI:** 10.64898/2025.12.08.25341479

**Authors:** Albert C. Ludolph, Jesus S. Mora, Patrick Vermersch, Alain Moussy, Colin D Mansfield, Olivier Hermine

## Abstract

**Background:** Amyotrophic lateral sclerosis (ALS) is a neurodegenerative disease that urgently requires new treatments. Masitinib, a tyrosine kinase inhibitor targeting microglia and mast cells, aims to slow disease progression by reducing neuroinflammation. A previously reported phase 2b/3 study showed that masitinib (4.5 mg/kg/day) with riluzole significantly slowed decline of the revised amyotrophic lateral sclerosis functional rating scale (ALSFRS-R) over 48 weeks in a specific patient population. However, a baseline imbalance was noted, with a higher proportion of patients with very severe loss of functionality (those with a score of 0 on at least one item of the ALSFRS-R) in the masitinib group. This post-hoc analysis evaluated the efficacy and safety of masitinib in the subgroup ‘ALS prior to any complete loss of functionality’ to eliminate potential bias and evaluate the effect of treatment in the early phase of the disease.

**Methods:** Data from study AB10015 were analyzed for patients with a score of at least 1 on all ALSFRS-R items at baseline. The primary endpoint was the change in ALSFRS-R score (ΔALSFRS-R) at week 48. Secondary endpoints included progression-free survival (PFS) and overall survival (OS).

**Results:** In the subgroup (N=84 for masitinib, N=104 for placebo), masitinib demonstrated an increase of the treatment effect compared with that in the primary analysis population. The ΔALSFRS-R difference between masitinib and placebo was 4.04 points (p=0.0065), an improvement from 3.39 points in the primary analysis. The median PFS gain increased from +4 months to a statistically significant +9 months (p=0.0057), and the median OS gain increased from +6 months to a statistically significant +12 months (p=0.0192). Safety was consistent with that of the overall study safety population or even improved, with the incidence of serious adverse events in the masitinib group decreasing from 27.6% to 22.6%.

**Conclusion:** Excluding patients with very severe loss of functionality eliminated baseline imbalances and revealed a favorable benefit-risk profile for masitinib treatment. The subgroup ‘ALS prior to any complete loss of functionality’ experienced consistently greater benefits in terms of function, survival, and quality of life. This subgroup is easily identifiable using the ALSFRS-R assessment and is an option as the target population for future development, informing the design of confirmatory study AB23005.

## Introduction

Amyotrophic lateral sclerosis (ALS) is a debilitating and life-threatening disease characterized by progressive loss of movement, respiratory insufficiency, and poor long-term survival. There is an urgent unmet medical need for new treatment options that can delay disease progression and/or enhance the quality of life of patients, particularly those with sporadic ALS.

Masitinib (AB1010) is a selective tyrosine kinase inhibitor (TKI) that inhibits colony-stimulating factor 1 receptor (CSF-1R), KIT, LYN, and FYN in the submicromolar range [Dubreuil 2009, Anastassiadis 2011, Davis 2011]. By targeting the CSF1/CSF-1R signaling pathway, masitinib regulates CSF-1R-dependent cells, such as microglia, which are known to play a pathogenic role in ALS progression [Brites 2014, Clarke 2020]. Furthermore, its action against KIT, LYN, and FYN enables masitinib to inhibit mast cell function and activation, which are key effector immune cells in chronic inflammatory processes [Krystel-Whittemore 2016, Angelini 2020]. Evidence also suggests that ALS involves interactions (cross-talk) between mast cells, microglia, and astrocytes, potentially resulting in motor neuron damage [Skaper 2018, Vahsen 2021, Sandhu 2021]. This multifaceted therapeutic approach aims to slow microglia-related disease progression, reduce neuroinflammation, and modulate the degenerative neuronal microenvironment in the central and peripheral nervous systems. The beneficial effects of masitinib treatment have been observed in preclinical animal studies [Kovacs 2021, Trias 2020, Harrison 2020, Trias 2018, Trias 2017, Trias 2016] and in humans through significant improvements in clinical measures of disease progression [Mora 2020, Mora 2021].

This article presents subgroup analyses of a previously published study on masitinib in ALS (AB10015, NCT02588677) [Mora 2020]. In brief, study AB10015 was an international, multicenter, phase 2b/3, randomized, double-blind, placebo-controlled trial that evaluated masitinib as an add-on therapy to riluzole (100 mg/day). The prespecified primary efficacy analysis population included patients with ALS with a progression rate from disease onset to baseline (ΔFS) of less than 1.1 points/month, as assessed by the revised amyotrophic lateral sclerosis functional rating scale (ALSFRS-R), and who received masitinib at 4.5 mg/kg/day (administered orally as two daily dosages). The primary endpoint was the change in the revised amyotrophic lateral sclerosis functional rating scale (ΔALSFRS-R) after 48 weeks of treatment. The results for masitinib 4.5 mg/kg/day indicated a significant benefit over placebo in ΔALSFRS-R, with a 3.39-point difference (p=0.016), representing a 27% decrease in ALSFRS-R decline over 48 weeks compared to placebo. This finding was confirmed by multiple sensitivity analyses. Additionally, a 25% delay in disease progression was observed based on progression-free survival (PFS) analysis (p=0.016). Other secondary endpoints demonstrated significant benefits in respiratory function and quality of life, including improvements in eating/drinking, daily activities, communication, and physical mobility. Overall, this study demonstrated that masitinib 4.5 mg/kg/day as an add-on to riluzole had a positive treatment effect with acceptable safety in ALS patients with ΔFS <1.1.

As elaborated below, a closer examination of the baseline characteristics revealed a notable imbalance in the distribution of ALSFRS-R scores between the treatment groups, even though the baseline total ALSFRS-R scores and average ALSFRS-R progression rates were well matched between the groups. The masitinib 4.5 mg/kg/day group had a higher proportion of patients with very severe loss of functionality, specifically those with at least one ALSFRS-R item score of zero, indicating an inability to perform certain tasks, than the placebo group. This imbalance occurred because, although the ALSFRS-R score was included among the key prognostic factors for minimization (a dynamic randomization algorithm designed to reduce any imbalance between treatment groups), there was no stratification by the ALSFRS-R severity category. This bias is important because masitinib is intended to slow disease progression rather than restore lost abilities. For diseases such as ALS, expecting a drug to recover lost functions is unrealistic, even if it can delay the progression. Therefore, administering masitinib before any complete loss of function aligns better with its intended mechanism, as it is expected to be most effective in patients who have not yet experienced a complete loss of functionality (i.e., a score of zero on any ALSFRS-R item). Consequently, it was crucial to assess the effect of masitinib on a cohort of patients with ALS who had not yet experienced any complete loss of function (i.e., a score of at least 1 on all 12 ALSFRS-R items) to eliminate potential bias.

Disease severity was categorized using the ALSFRS-R, a validated rating instrument for monitoring the progression of disability in patients with ALS. The ALSFRS-R measures 12 aspects of physical function, organized into four domains, to assess bulbar, fine motor, gross motor, and respiratory symptoms. To quantify the loss of functionality in an ordinal manner, the ALSFRS-R anchor points rate functionality from 4 (normal) to 0 (no ability) for each item, with a maximum total score of 48 and a minimum score of 0. Thus, severity can be defined in terms of the remaining functionality of the ALSFRS-R items or the global ALSFRS-R score. A serious drawback of the latter approach is its assumption of uniform decline across all items, which fails to adequately account for the complete loss of functionality that may have occurred in one item, even if global functionality remains relatively high (a consequence of the scale’s multidimensionality). Therefore, the following categories of ALSFRS-R severity were defined:

- Very severe loss of functionality: Patients had a score of 0 (i.e., complete loss of function) on at least one individual item of the ALSFRS-R scale.
- Severe loss of functionality: Patients had a score of 1 (i.e., severe impairment of function) for at least one of the individual ALSFRS-R items and a score greater than zero for all other items.
- Moderate (or non-severe) loss of functionality: Patients had a score of 2 (i.e., moderate impairment of function) for at least one of the individual ALSFRS-R items and a score greater than 1 for all other items.
- Mild loss of functionality: Patients had a score of 3 (i.e., mild impairment of function) for at least one of the individual ALSFRS-R items and a score greater than 2 for all other items.

The subgroup ‘ALS prior to any complete loss of functionality’ is thus clearly defined and clinically relevant, with precedents in the literature for such ALSFRS-R severity-based patient categorization [Takahashi 2017, Writing Group 2017, Imamura 2024].

## Methods

Details regarding the method and patient selection criteria of study AB10015 have been published previously [Mora 2020]. Briefly, patient randomization was conducted using a minimization method based on baseline covariates, including the site of onset (spinal versus bulbar), ALSFRS-R score, age, geographical region, and ΔFS. Patients receiving masitinib 4.5 mg/kg/day with a post-onset ΔFS of less than 1.1 points/month were designated as the primary efficacy analysis population. Eligible patients were aged 18–75 years with a laboratory-supported probable, probable, or definite diagnosis of ALS, a disease duration of less than 36 months from the first ALS symptom, and forced vital capacity (FVC) of at least 60% at baseline. Additionally, patients were required to be on a stable dose of riluzole (100 mg/day) for at least 30 days prior to baseline. All patients provided written informed consent, which included permission to review their medical records to complete undetermined endpoints, such as overall survival.

The primary endpoint was a decline in the ALSFRS-R score at week 48 (ΔALSFRS-R), with any patient who died after randomization having an imputed ALSFRS-R score of zero. ΔALSFRS-R was calculated using an analysis of covariance model adjusted for baseline covariates, and the results were expressed as the difference in least square means (ΔLSM) between treatments (masitinib versus placebo). Secondary endpoints included the change from baseline in the ALS Assessment Questionnaire 40-item (ALSAQ-40) score, change from baseline in FVC, overall survival (OS), time-to-event analysis (an endpoint driven by both death and fixed disease progression on the ALSFRS-R scale), defined as a decline of nine points from baseline or death, and the Combined Assessment of Function and Survival (CAFS). Overall survival was defined as the time from randomization to death from any cause, with relevant placebo patients censored at the time of switching to masitinib treatment as part of an international early access compassionate use program. Overall survival p-values were calculated using the multivariate log-rank test with the aforementioned covariates.

The AB10015 protocol specifies that missing data should be imputed using the last observation carried forward (LOCF) method. For this subgroup analysis, we also employed the more robust copy increments in reference (CIR) methodology, which is the most context□specific and appropriate imputation approach. This is because participants are expected to maintain the treatment benefits accrued until dropout, after which they follow the outcome trajectory of the reference group [Leurent 2020].

## Results

### Baseline imbalance in ALS severity between treatment groups

Further examination of the potential baseline confounders revealed an imbalance between the treatment groups in terms of the distribution of patients with very severe loss of functionality. A larger proportion of patients with ALS who had completely lost function in at least one component of the ALSFRS-R (i.e., score of zero on any item) were assigned to the masitinib 4.5 mg/kg/day treatment arm (21/105, 20%) than to the control arm (9/113, 8%) (Table 1). Additionally, within this subset, disease severity was more pronounced in the masitinib group, with more items scored as zero, indicating a worse functional status. Specifically, the masitinib arm included 21 patients with 37 zero scores, considerably surpassing the placebo group of 9 patients with 12 zero scores (Table 2).

**Table 1:** Distribution of patients with very severe ALS in the primary analysis population of study AB10015.

|  | Placebo | Masitinib 4.5 |
| --- | --- | --- |
| **Primary analysis population; n (%)** | 113 (100) | 105 (100) |
| **Subgroup\*; n (%)** | 104 (92) | 84 (80) |
| **Patients excluded from subgroup\*\*; n (%)** | 9 (8) | 21 (20) |

\* ALS prior to any complete loss of function. \*\* Patients with very severe loss of functionality. Masitinib 4.5 = masitinib 4.5 mg/kg/day treatment arm.

**Table 2:** Number of zero scoring items (very severe functional loss) in the primary analysis population of study AB10015.

|  | Placebo | Masitinib 4.5 |
| --- | --- | --- |
| 1 item with zero score; n [Q] | 8 [8] | 10 [10] |
| 2 items with zero score; n [Q] | 0 | 7 [14] |
| 3 items with zero score; n [Q] | 0 | 3 [9] |
| 4 items with zero score; n [Q] | 1 [4] | 1 [4] |
| **Total items with zero score; n [Q]** | 9 [12] | 21 [37] |

[Q] Represents the number of items with zero score. Some patients (n) had a zero score (Q) for more than one item. Masitinib 4.5 = masitinib 4.5 mg/kg/day treatment arm.

### Baseline characteristics of the subgroup ‘ALS prior to any complete loss of functionality’

After excluding all patients with very severe functional impairment, the remaining subgroup comprised 85% of the AB10015 primary analysis population. The baseline characteristics of the placebo group (N=104) and the masitinib 4.5 mg/kg/day treatment group (N=84) remained well-matched across several key demographic, clinical, and geographic variables (Table 3). For example, the baseline ALSFRS-R scores for the placebo and masitinib groups were nearly identical, averaging 39.8 ± 4.3 and 39.9 ± 4.2, respectively. The average monthly decline in ΔFS was also comparable between the groups, with means of 0.49 ± 0.25 for placebo and 0.45 ± 0.25 for masitinib-treated patients. The age distributions were similar, with mean ages of 55.4 ± 10.8 years and 54.3 ± 10.7 years, respectively. Pulmonary function, measured by forced vital capacity (FVC) as a percentage of the predicted value, showed mean values of 90.9 ± 18.4% for placebo and 91.8 ± 16.0% for the masitinib group. Regarding the site of disease onset, spinal onset was predominant in both groups (77.9% placebo and 81.0% masitinib). These comparable demographic, clinical, and regional data confirmed the balanced baseline characteristics between the treatment arms, supporting the validity of the comparative analyses.

**Table 3:**
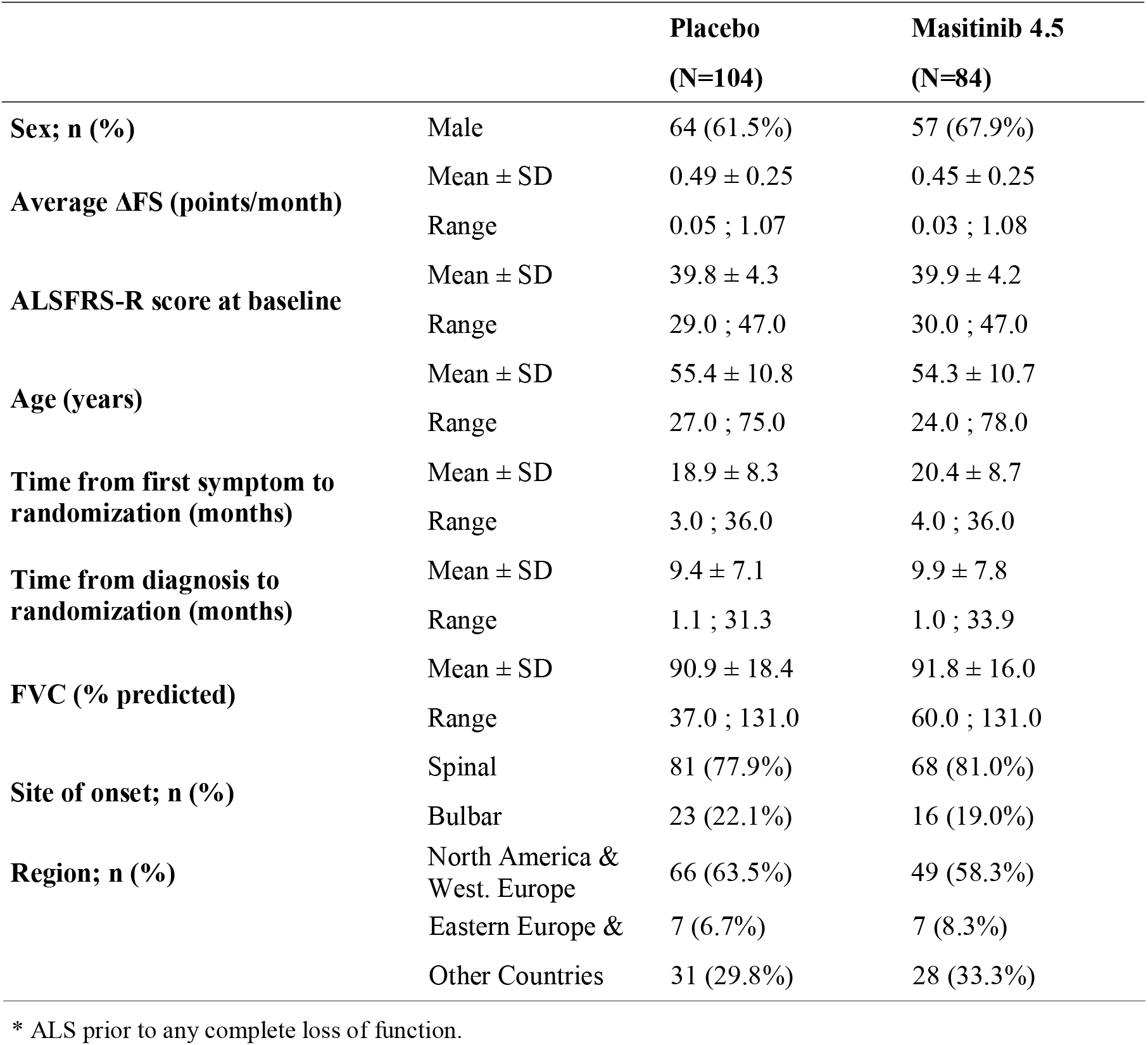
Subgroup* baseline characteristics.

### Efficacy in the subgroup ‘ALS prior to any complete loss of functionality’

Results from the subgroup analysis showed an increase in treatment effect across multiple efficacy endpoints compared with the primary analysis population of study AB10015 (Table 4). Specifically, the predefined primary endpoint (ΔALSFRS-R, LOCF analysis) showed a significant difference of least squares means between treatment arms (masitinib versus placebo) of 4.04 (p=0.0065) in the subgroup compared with 3.39 (p=0.0157) in the primary analysis population. Analysis using the more appropriate CIR methodology maintained this significant treatment effect, with a between-group difference of 3.13 (p=0.0308) in the subgroup compared with 2.68 (p=0.0462) in the primary population. The Combined Assessment of Function and Survival (CAFS) indicated a relative benefit increase from +14.8% in the primary analysis population to +20.2% in the subgroup, with the latter reaching statistical significance (p=0.0290). Similarly, the benefit in respiratory function, as measured by FVC (CIR methodology), increased from 5.85 in the primary analysis population to 7.59 in the subgroup, with the latter reaching statistical significance (p=0.00384). Patient-reported quality of life, as measured by the ALSAQ-40 (CIR methodology), showed a slight improvement in the subgroup (-6.22, p=0.044) compared with that in the primary analysis population (-6.04, p = 0.0305).

**Table 4:** Masitinib treatment effect, with respect to placebo, in the subgroup* compared to the primary analysis population of study AB10015.

|  |  | **Primary analysis**<br>**population** | **Subgroup** |
| --- | --- | --- | --- |
| **ΔALSFRS-R (mLOCF)**<br>**(primary analysis)** | Diff. of mean | 3.39 | 4.04 |
|  | p-value | 0.0157 | 0.0065 |
| **ΔALSFRS-R (CIR)** | Diff. of mean | 2.68 | 3.13 |
|  | p-value | 0.0462 | 0.0308 |
| **CAFS** | Relative benefit | + 14.8% | + 20.2% |
|  | p-value | 0.0776 | 0.0290 |
| **ALSAQ-40 (CIR)** | Diff. of mean | -6.04 [-11.51;-0.57] | -6.22 [-12.27;-0.17] |
|  | p-value | 0.0305 | 0.044 |
| **FVC (CIR)** | Diff. of mean | 5.85 [-0.98;12.67] | 7.59 [0.41;14.77] |
|  | p-value | 0.0931 | 0.0384 |

\* ALS prior to any complete loss of functionality

### Survival analyses in the subgroup ‘ALS prior to any complete loss of functionality’

Although no formal regulatory threshold has been established, the ALS therapeutic community may regard a median survival gain of approximately 4–6 months (or preferably more) as clinically meaningful, especially when accompanied by improvements in function or quality of life [Khairoalsindi 2018, Nikitin 2023].

Masitinib use resulted in a statistically significant and clinically relevant improvement in functional progression, as assessed by median PFS. This increased from +4 months (p=0.0159) relative to placebo in the primary analysis population to +9 months (p=0.0057) in the subgroup, with the latter achieving statistical significance (Table 5 and Figure 1A).

**Table 5:** Masitinib survival benefit in the subgroup* compared to the primary analysis population of study AB10015.

|  |  | Primary analysis population | Subgroup* |
| --- | --- | --- | --- |
| **Median PFS** | Gain (vs placebo) | + 4 months | + 9 months |
|  | Median [95% CI] | 20 [14; 30] vs 16 [11; 19] | 25 [17, NE] vs 16 [11, 19] |
|  | p-value log rank | 0.0159 | 0.0057 |
| **Median OS (Long-term)** | Gain (vs placebo) | + 6 months | + 12 months |
|  | Median [95% CI] | 46 [33; 69] vs 40 [30; 49] | 53 [36; NE] vs 41 [30; 54] |
|  | p-value log rank | 0.0761 | 0.0192 |

\* ALS prior to any complete loss of functionality

**Figure 1.**
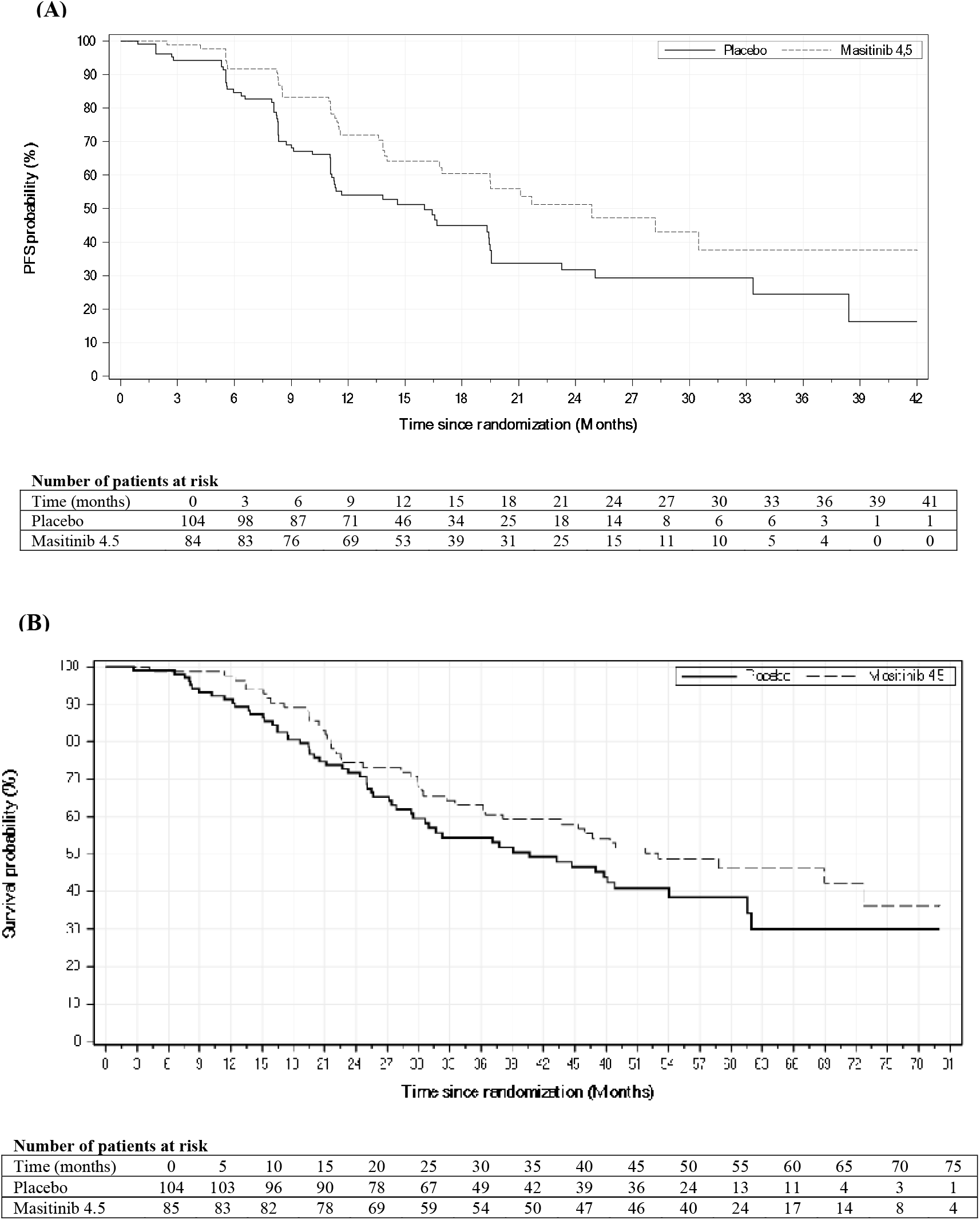
Subgroup ‘ALS prior to any complete loss of functionality’ (A) Kaplan–Meier analysis of PFS. (B) Kaplan–Meier analysis of OS (long-term follow-up).

Regarding long-term OS, the median OS increased from +6 months (relative to placebo) in the primary analysis population to +12 months (p=0.0192) in the subgroup, with the latter reaching statistical significance (Table 5 and Figure 1B).

### Safety in subgroup ‘ALS prior to any complete loss of functionality’

The safety profile of masitinib in the subgroup showed notable improvements compared to the primary analysis safety population (Table 6). Specifically, the incidence of serious adverse events (AEs) in patients receiving masitinib decreased from 27.6% in the primary population to 22.6% in the subgroup (compared with 16.7% and 16.3% in the placebo arm, respectively). The incidence of AE (all grades), severe AEs, and death related to AE was also slightly reduced in the subgroup compared with the primary analysis safety population. Overall, masitinib treatment was associated with a higher frequency of adverse events than placebo; however, patients in this subgroup had a better safety profile with masitinib than those in the primary analysis safety population.

**Table 6:**
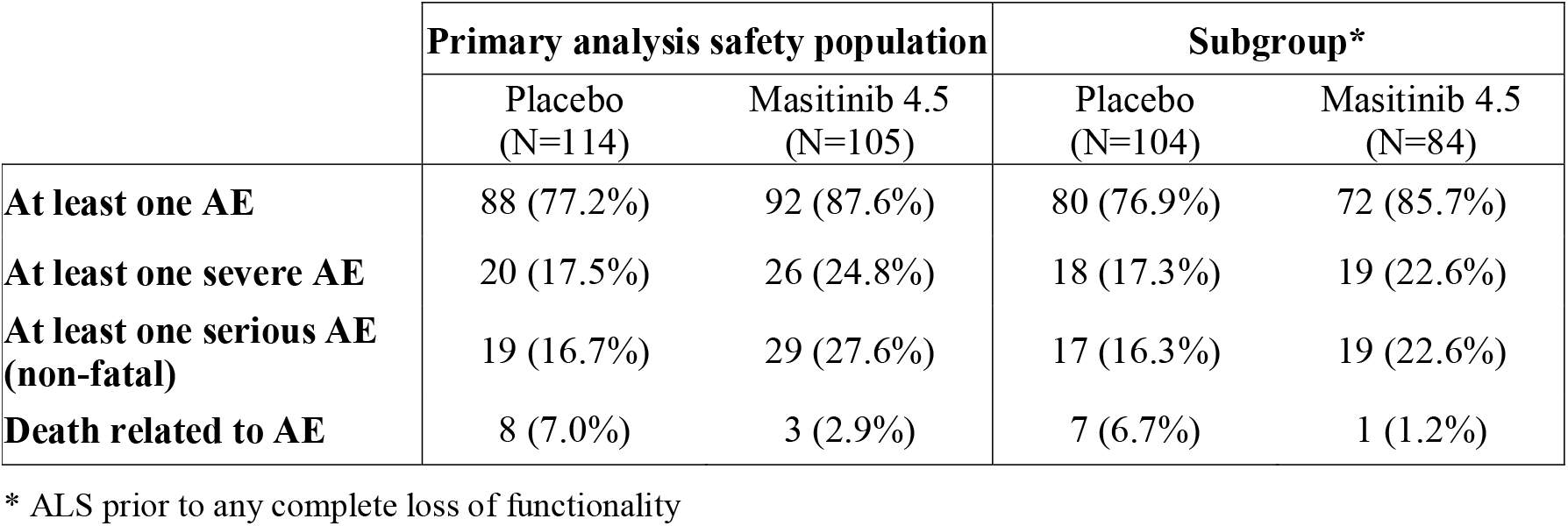
Differential safety overview between the subgroup* and the primary analysis safety population of study AB10015.

## Discussion

When the primary endpoint of a randomized clinical trial shows a significant benefit, as was the case for the primary analysis population of study AB10015 [Mora 2020], it is methodologically sound to ensure that the conclusions are consistently applicable across subgroups of the trial population, or alternatively, to identify post-hoc a subgroup that optimizes the benefit-risk balance [EMA 2019]. A baseline imbalance in ALS severity distribution was observed in the primary analysis population, with a higher number of patients with very severe loss of functionality assigned to the masitinib 4.5 mg/kg/day group than to the control group, which may have adversely affected the measured treatment effect. This bias was eliminated by excluding these patients (i.e., those scoring zero on at least one item of the ALSFRS-R), and the subsequent subgroup evaluation provided corroboratory evidence for the positive treatment effect of masitinib in ALS. However, because this was an unplanned post-hoc analysis without adjustment for multiplicity, the results should be considered hypothesis-generating.

Importantly, there is a biological plausibility for the differential effect of treatment in the defined subgroups. The action of masitinib in ALS has been thoroughly demonstrated using a relevant neurodegenerative model that mirrors the complexity of a multicomponent immune response [Kovacs 2021, Trias 2020, Harrison 2020, Trias 2018, Trias 2017, Trias 2016]. The findings indicate that instead of regenerating motor neurons, masitinib can reduce the rate of motor neuron death, thereby slowing disease progression by modulating mast cell and microglial activity, reducing mast cell, macrophage, and neutrophil infiltration, preventing terminal Schwann cell loss, and enhancing reinnervation in partially denervated plantaris muscles. Overall, this provides a credible mechanistic explanation for the more pronounced treatment effect when masitinib was initiated at a less severe stage of the disease, while more motoneurons were still functioning.

Considering the clinical feasibility of identifying the subgroup ‘ALS prior to any complete loss of functionality’, the ALSFRS-R stands out as a validated rating instrument for assessing disease severity in ALS. This metric is routinely evaluated and documented in clinical practice, as evidenced by its inclusion in the ALS Toolkit [Granit 2022] and the European Network to Cure ALS (ENCALS) survival model [Westeneng 2018]. Finally, Ludolph et al. emphasized the importance of the ALSFRS-R in routine management of ALS and its widespread application beyond clinical trials. The authors concluded that because the ALSFRS-R is derived from information collected during standard patient care and monitoring, it serves as an effective tool for patient selection by treating physicians, with no barriers to its use in clinical practice [Ludolph 2024].

No treatment has been shown to effectively halt or significantly reverse ALS progression. Modest survival benefits [Miller 2012], inconsistencies in post-marketing efficacy data [Gupta 2025, Kashyap 2025, Takahashi 2025, Shefner 2024, Witzel 2022, Brooks 2022, Ortiz 2020, Lunetta 2020, Fortuna 2019], and a shift towards more targeted therapies for specific genetic subtypes of ALS [Miller 2022] highlight the ongoing challenges in developing broadly effective treatments for ALS and underscore the urgent need for novel therapies and patient enrichment strategies, which this subgroup analysis of masitinib aims to address.

## Conclusion

Excluding patients with very severe loss of functionality from the dataset of study AB10015 resulted in balanced baseline characteristics between the treatment arms and eliminated a potential source of bias that may have obscured the true potential of masitinib. The resulting subgroup showed consistent improvements across efficacy endpoints, including survival, and safety profile enhancements. The subgroup descriptor ‘ALS prior to any complete loss of functionality’ is practical for clinical use, as it is based on the established ALSFRS-R assessment. Moreover, this aligns well with the mechanism of action of masitinib and the therapeutic goal of preserving neuromuscular function rather than repairing existing neurological damage.

Overall, the subgroup experienced consistently greater and statistically more robust benefits from treatment in terms of functionality, survival, and quality of life than the broader primary analysis population. Therefore, such a subgroup might be useful for the evaluation of the masitinib effect, accompanied by emerging biomarkers sensitive to inflammation. Consequently, the benefit-risk balance is more favorable in this subgroup, making it the recommended patient population for future clinical development of masitinib for ALS. Indeed, the findings from this analysis have been integrated into the design of the confirmatory masitinib study AB23005 (NCT07174492), thereby targeting a population that optimizes the benefit-risk balance to enhance the likelihood of study success and achieving a clinically relevant beneficial effect.

## Data Availability

All data produced in the present study are available upon reasonable request to the authors

